# White matter tracts associated with iTBS-induced heart rate deceleration and treatment response in major depressive disorder

**DOI:** 10.1101/2025.01.09.25320274

**Authors:** Jonas Wilkening, Roberto Goya-Maldonado

**Affiliations:** Laboratory of Systems Neuroscience and Imaging in Psychiatry (SNIP-Lab), Department of Psychiatry and Psychotherapy, University Medical Center Göttingen, Göttingen, Germany

## Abstract

Intermittent theta burst stimulation (iTBS) is a well-established treatment for major depressive disorder (MDD), but predicting clinical outcomes remains challenging. Heart rate modulation induced by iTBS has emerged as a potential biomarker for treatment response, yet the role of white matter (WM) properties in mediating these effects is largely unexplored. In this quadruple-blind, crossover study, we investigated the relationship between WM microstructure, iTBS-driven heart rate deceleration, and antidepressant effects. Using correlational tractography, we analyzed four major WM tracts—the cingulum, fornix, superior longitudinal fasciculus, and uncinate fasciculus—and examined short-term WM microstructural changes to assess their predictive value for therapeutic outcomes. Baseline WM findings revealed that in the fornix and right dorsal cingulum fractional anisotropy (FA) negatively correlated with heart rate deceleration. Radial and mean diffusivity (MD, RD) in the fornix positively correlated with heart rate deceleration. FA in the right ventral cingulum positively correlated while MD and RD negatively correlated with symptom improvement. Longitudinally, FA increases in the left cingulum were significantly associated with greater symptom alleviation after treatment. Notably, iTBS-induced heart rate modulations correlated with clinical improvement after six weeks, while WM microstructural properties in the fornix and cingulum demonstrated predictive value for both heart rate modulation and treatment response. WM changes in the cingulum, evident as early as four weeks, highlight its unique neuroplasticity potential along iTBS intervention. Together, these findings provide novel insights into the structural connectivity patterns influencing iTBS outcomes, offering a novel foundation for more personalized therapeutic strategies in MDD.

## 1. Introduction

Major depressive disorder (MDD) is a pervasive mental health condition with significant societal and individual impacts, marked by rising prevalence globally (World Health Organization, 2017). While pharmacological and psychological interventions remain central to treatment, novel neuromodulatory approaches, such as intermittent theta burst stimulation (iTBS), have garnered attention for their potential efficacy in alleviating depressive symptoms. iTBS, a patterned form of repetitive transcranial magnetic stimulation (rTMS), applies rapid bursts of stimulation to the left dorsolateral prefrontal cortex (DLPFC). This technique has been shown to reduce depressive symptoms, offering promise as an alternative or adjunctive therapy for patients who do not respond to conventional treatments (Blumberger et al., 2018; Cole et al., 2022). However, predicting which individuals will benefit from iTBS remains a significant challenge, and identifying reliable biomarkers of treatment response is an ongoing area of research.

Among these potential biomarkers, heart rate (HR) modulation during iTBS has emerged as a promising indicator of therapeutic outcomes. Individuals with MDD frequently exhibit autonomic nervous system dysregulation, evidenced by increased resting HR and reduced heart rate variability (HRV) compared to healthy individuals (Kidwell & Ellenbroek, 2018; Koch et al., 2019; Licht et al., 2008). This autonomic imbalance reflects altered interactions between the brain and peripheral systems, particularly within the central autonomic network (CAN). Notably, high-frequency rTMS targeting the DLPFC has been shown to acutely reduce HR, a change not observed with sham stimulation (Dijkstra et al., 2024; Iseger et al., 2021; Udupa et al., 2007). Furthermore, HR and HRV parameters changes along stimulation have been correlated with reductions in symptom severity (Cosmo et al., 2022; Iseger et al., 2020). These findings suggest that HR modulation may serve as a functional biomarker of iTBS efficacy, reflecting changes in the frontal-vagal network that integrates autonomic and cognitive processes (Iseger et al., 2020). While this supports a role for functional mechanisms, the structural pathways underpinning these effects remain poorly understood.

White matter (WM) microstructure may play a pivotal role in mediating the effects of iTBS on both HR modulation and depressive symptoms. Altered WM integrity is a well-documented feature of MDD, with diffusion tensor imaging (DTI) studies revealing widespread microstructural abnormalities in individuals with the disorder compared to healthy controls (Bracht et al., 2015; Olvet et al., 2016; van Velzen et al., 2020). Fractional anisotropy (FA), a DTI-derived metric of WM integrity, has been implicated as a potential predictor of treatment response. For example, prior research has shown that higher FA values in specific WM tracts are associated with clinical improvement following antidepressant therapies, including pharmacological and neuromodulatory interventions (Barredo et al., 2019; Klooster et al., 2020; Korgaonkar et al., 2014; Ning et al., 2022). Importantly, these findings suggest that WM pathways may provide insight into the brain’s capacity for neuroplasticity, which could be essential for understanding iTBS efficacy. However, the specific WM tracts associated with iTBS-induced changes in HR and symptom alleviation have yet to be systematically investigated.

Beyond baseline WM properties, longitudinal changes in WM microstructure may also hold predictive value for treatment outcomes. Neuromodulatory interventions, including rTMS, have been shown to induce rapid changes in WM integrity. For instance, increases in FA and reductions in mean diffusivity (MD) and radial diffusivity (RD) have been observed as early as one week after treatment, correlating with clinical improvement in mood disorders (Hofstetter et al., 2013; Ning et al., 2022; Tura et al., 2024). These findings suggest that WM reorganization may reflect dynamic neuroplastic processes triggered by iTBS. Crucially, such changes are not limited to the stimulation site but extend to broader regions, highlighting the widespread impact of neuromodulation on brain connectivity. By combining baseline and longitudinal analyses, it may be possible to capture both stable WM characteristics that predispose individuals to treatment response and dynamic changes that signal neuroplasticity during therapy.

To investigate these mechanisms, advanced imaging techniques can provide greater precision in assessing WM microstructure. For example, free-water correction enhances the specificity of diffusion metrics by separating extracellular diffusion components, such as edema, from anisotropic diffusion within axons (Pasternak et al., 2009). Connectometry, a complementary method, maps structural connectivity patterns linked to study variables with higher sensitivity than traditional tractography(Yeh et al., 2016). These methods address limitations associated with traditional DTI, such as partial volume effects, enabling more accurate identification of WM pathways associated with treatment outcomes.

Several WM tracts stand out as candidates for mediating the effects of iTBS in MDD. The cingulum, which interconnects regions of the default mode network (DMN) and the limbic system, plays a critical role in emotional regulation and reward processing (Weigand et al., 2018). Greater WM integrity in the cingulum has been linked to antidepressant response, possibly due to its role in modulating activity in the subgenual anterior cingulate cortex (sgACC), a key region implicated in MDD pathology and HR control (Dijkstra et al., 2024; Fox et al., 2012; Liston et al., 2014). Similarly, the fornix, a major output tract of the hippocampus that connects to the hypothalamus and amygdala, is integral to memory and autonomic regulation. FA values in the fornix have been associated with emotional regulation and autonomic function, suggesting its relevance to iTBS-driven HR modulation and clinical outcomes(Grieve et al., 2016; Thayer & Lane, 2000).

In addition, the superior longitudinal fasciculus (SLF) and uncinate fasciculus have been implicated in MDD and its treatment. The SLF, which connects the DLPFC to parietal regions involved in attention and executive function, has been associated with treatment resistance, with lower FA values linked to reduced responsiveness to antidepressant therapies (Murphy & Frodl, 2011). Meanwhile, the uncinate fasciculus, a tract connecting the sgACC to the amygdala, plays a key role in emotion regulation. This tract has been associated with clinical improvement following deep brain stimulation, highlighting its potential role in mediating therapeutic effects in MDD (Liu et al., 2016; Riva-Posse et al., 2014). Together, these four tracts form an interconnected network that integrates cognitive, emotional, and autonomic functions, making them prime candidates for understanding the mechanisms underlying iTBS efficacy.

In the present study, we examined the relationship between WM microstructure, HR modulation, and clinical improvement in MDD, focusing on four major tracts anatomically relevant to the iTBS stimulation site: the cingulum, fornix, SLF, and uncinate fasciculus. Our dual approach combined baseline and longitudinal analyses to provide a comprehensive understanding of how WM properties relate to iTBS efficacy. We hypothesized that tracts with higher baseline microstructural integrity, as reflected by higher FA as well as lower MD and RD, would predict greater HR deceleration and stronger symptom alleviation. Additionally, we expected to observe longitudinal increases in FA next to decreases in MD and RD in these tracts, with these changes correlating positively with clinical improvement. By integrating functional biomarkers like HR modulation with WM structural metrics, this study aims to advance our understanding of the mechanisms underlying iTBS efficacy and pave the way for more personalized therapeutic strategies.

## 2. Materials and Methods

### Participants

The study protocol (clinicaltrials.gov/show/NCT05260086) is accordance with the latest version of the Declaration of Helsinki and was approved by the Ethics Committee of the University Medical Center Göttingen (UMG). Prior to enrollment, all participants provided verbal and written informed consent after the study protocol was fully explained. Individuals aged 18 to 60 years, diagnosed with MDD and currently experiencing a moderate or severe depressive episode, were recruited for the study. Trained psychiatrists confirmed the inclusion criteria using the Structured Clinical Interview for DSM-5 Disorders - Clinical Version (SCID-V-CV) and the the Montgomery-Åsberg Depression Rating Scale (MADRS) (Hengartner et al., 2020; Montgomery & Asberg, 1979).

Patients with contraindications to MRI or rTMS (e.g., epilepsy, neurological diseases, pregnancy, metallic implants) were excluded from the study. Participants were required to maintain stable medication regimens for a minimum of two weeks prior to and throughout the duration of the study, with adherence monitored through serum level assessments. These values were also incorporated into the statistical model to account for their potential effects on clinical improverement. The sample included both clinic inpatients and outpatients; however, the study intervention was administered exclusively in an outpatient setting. Detailed study information can be found elsewhere (Wilkening et al., 2022).

### Study design

The study protocol spanned six weeks and employed a quadruple-blinded (participant, care provider, investigator, and rater) randomized sham-controlled crossover design (Fig. 1). Each participant received one week of active stimulation and one week of sham stimulation, with the order of conditions randomly assigned (active-sham, sham-active). Randomization followed pre-coded sequences generated using a true random function in MATLAB (The MathWorks, Inc., Natick, MA, USA) and was managed by the study principal investigator, who had no direct contact with participants.

**Figure 1:**
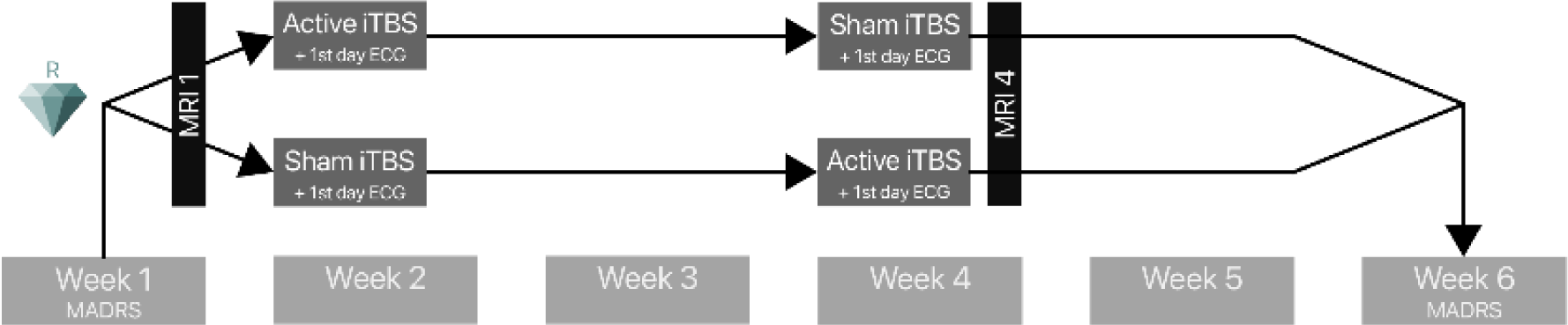
Study Design. The acelerated iTBS stimulation protocol was administered in a week over five consecutive days, before or after an identical sham protocol, in a crossover design. The MADRS score was evaluated at baseline and at a six-week follow-up, yielding a delta MADRS score. MRI scans were acquired before and after iTBS intervention. ECG recordings were obtained on the first day of each intervention week. iTBS: Intermittent theta burst stimulation, MADRS: Montgomery-Åsberg depression rating scale, ECG: Electrocardiogram.

As part of a broader investigation into the clinical benefit of personalized stimulation sites, an additional pseudorandomization was performed to compare two stimulation approaches: fixed stimulation at the EEG 10-20 cap F3 position or a personalized approach. The personalized approach involved selecting a point within the left DLPFC in which the individual exhibited the strongest anticorrelation with the DMN as measured by resting-state functional MRI (Singh et al., 2020; Wilkening et al., in preparation). In this study, we did not consider this separation, opting for a unified stimulation approach due to the similarity of the stimulation methods.

A nine-day gap separated the two stimulation weeks. Trained clinical interviewers collected the MADRS scores at the end of each week, starting from baseline, to assess the clinical outcome over the six-week period (Fig. 1).

### iTBS treatment

The MagVenture X100 system with Mag-option and a figure-of-eight MCF-B65 A/P cooled coil was used for stimulation by trained and experienced personnel. Each daily visit included four sessions of approximately 10 minutes each, followed by at least 20-minute pauses, based the necessary inter-session time described on previous literature (Tse et al., 2018). The iTBS protocol consisted of 5 Hz bursts comprising 3 pulses at 50 Hz, resulting in a volley with 2 seconds on and 8 seconds off. One session consisted of 60 volleys, resulting in a total of 1800 pulses per session, 7200 per day, 36000 pulses in total. The resting motor threshold (RMT) was reevaluated daily via electromyography in the right first dorsal interosseous muscle with the lowest intensity eliciting 50 mV motor reaction in 5 of 10 attempts. Treatment stimulation was performed at 110% of the RMT. In the sham condition, the same figure-eight-shaped MCF-B65 A/P coil was blindly flipped 180 degrees. Additionally, tENS electrodes mimicked scalp sensation simultaneously to the sound of magnetic pulses, regardless of the stimulation condition. The blinding procedure was assessed using a visual analogue scale.

### Cardiac data acquisition and analysis

During iTBS treatment, subjects had an ECG recorded using three chest electrodes from the NCG-rTMS device (neuroConn NCG-rTMS device, neuroCare, Munich, Germany), allowing synchronization with the triggers of the TMS coil. The ECG signal underwent powerline noise reduction using the SciPy module in Python 3.6 to and BioSignal Processing (BioSPPy) to remove band wander noise (Virtanen et al., 2020). The automatically detected R-peaks were then corrected to match with the maxima of the curve within a tolerance of 50 milliseconds. Subsequently, the RR intervals were calculated using a Python implementation of the algorithm developed by Lipponen & Tarvainen, 2019, allowing correction for ectopic beats. The slopes of RR intervals were calculated for 45 s after iTBS session initiation, which is the timeframe with the strongest difference between active and sham stimulation in Iseger et al., 2020. A positive slope indicates an increase in RR intervals, i.e. a decrease in HR. Outliers greater than ±2.5 in z-standardized values were removed on a session-wise basis. The median HR slope of all four sessions of the first day of stimulation was used as a predictive parameter (Iseger et al., 2017).

It was previously demonstrated that the change in the slope of RR intervals in the first 45 seconds of stimulation was significantly associated with the six-week treatment outcome (Suppl. Fig. 2A).

### Image acquisition

We analyzed MRI data from participants before the stimulation and after the second week of stimulation, resulting in an interval of 4 weeks (Fig. 1). All scans were acquired on a 3T Siemens scanner (Magnetom Prisma Fit, Siemens Healthcare, Erlangen, Germany) using a 32-channel head coil. The diffusion MRI was acquired at b=1000 s/mm^2^ with 64 directions including the b0 image, TR = 4200ms, TE = 93ms, 90 slices, Flip angle = 90°, FOV = 218mm, multiband factor = 3, fat saturation enabled. The voxel size was 1.7mm isotropic.

### Image processing

In a total of 34 DTI scans, one slice of one of the 64 volumes with diffusion weighting had to be removed because of artifacts using an in-house-written MATLAB script. After correction of diffusion data for eddy current and motion artifacts using FSL (Andersson & Sotiropoulos, 2016) the rotated bvecs were used for subsequent analysis (Jenkinson et al., 2012; Leemans & Jones, 2009) (Fig. 2A).

**Figure 2:**
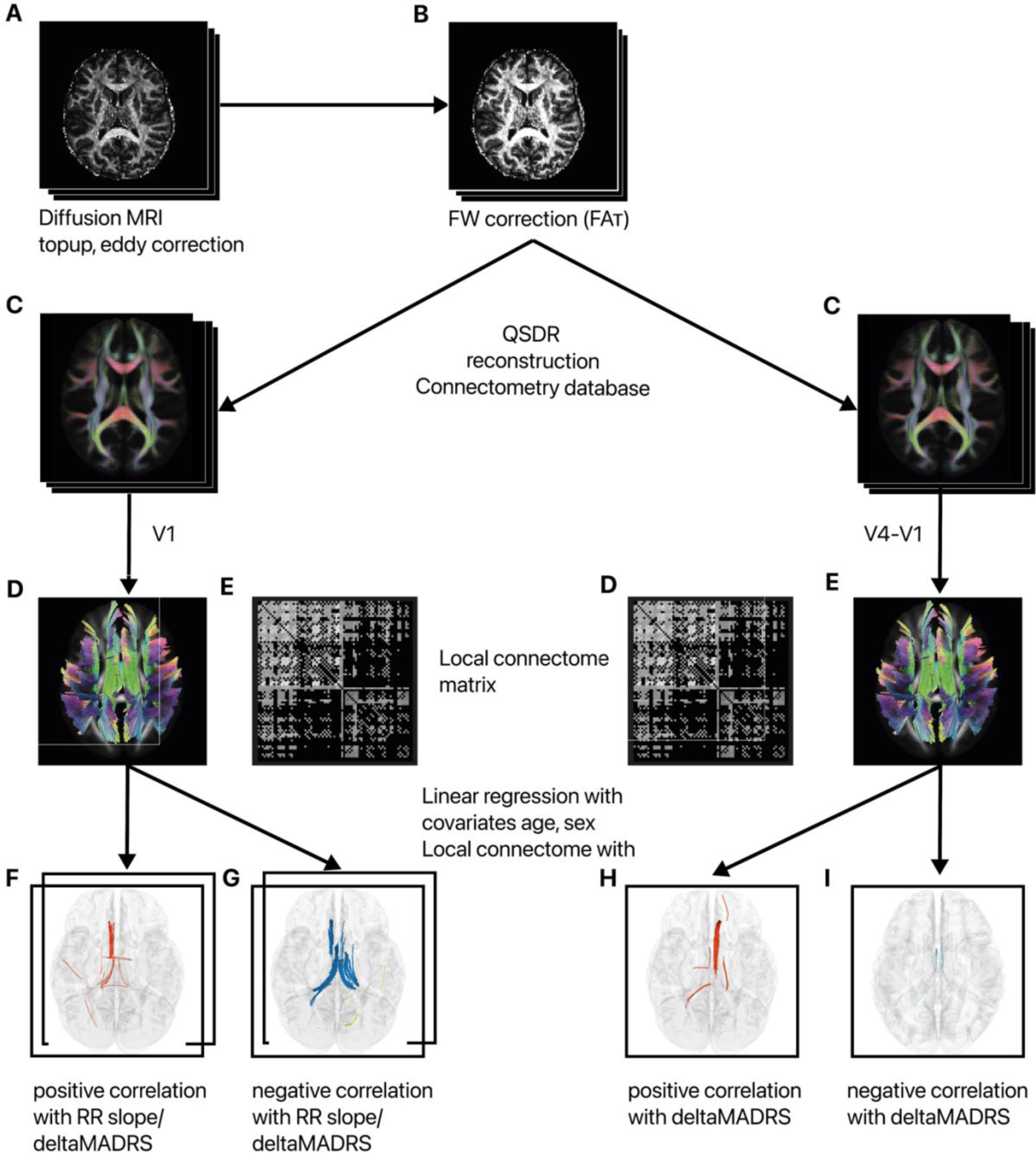
Diffusion imaging analysis pipeline. The diffusion analysis following the standard FSL processing pipeline, incorporating a FW-corrected protocol for DTI data. Further processing was conducted using DSI Studio’s QSDR reconstruction to generate a connectometry database for baseline data and a subtraction database for longitudinal correlational tractography. Key study variables included delta MADRS scores and HR modulations, represented by the RR-slope. FW: Free-water, Q-Space Diffeomorphic Reconstruction, MADRS: Montgomery-Åsberg depression rating scale, HR: heart rate, RR: Time between two successive ventricular depolarizations.

The data was input into a custom modified Python code (https://github.com/sameerd/DiffusionTensorImaging) referring to Pasternak et al., 2009. It describes a voxel-wise bi-tensor model which reflect the partial volume artifacts by free water and the tissue compartment (Hoy et al., 2014; Pasternak et al., 2009). The corrected images were fit with FA_T_, MD_T,_ RD_T_ using the dipy.reconstruct.dti function in Python 3.6 (Fig. 2B).

All subsequent analyses were conducted using DSI Studio (version 2022.09.27), following standard analysis protocols. The corrected images were reformatted into a DSI Studio-compatible format and resampled to an isotropic resolution of 2 mm. Integrity and quality checks were performed automatically (Yeh, Zaydan, et al., 2019) and visually inspected.

The diffusion data was reconstructed in the MNI space using q-space diffeomorphic reconstruction (QSDR) (Yeh & Tseng, 2011) (Fig. 2C) to obtain the spin distribution function (Yeh et al., 2010). The default diffusion sampling length ratio was 1.25. A subject-specific brain mask from FSL tool bet was applied (Smith, 2002), and Gaussian filtering (Fadnavis et al., 2020) was performed on the DWI signals using DSI Studio to achieve the necessary signal-to-noise ratio for correlational tractography, based on consultation and data inspection provided by the DSI Studio developer. FA_T_, MD_T,_ and RD_T_ maps were registered to the MNI space using QSDR.

### Connectometry

From the preprocessed data, a connectometry database file for both baseline and longitudinal analysis was created. In the latter case, the algorithm created a subtraction image (V4-V1).

Diffusion MRI connectometry (Yeh et al., 2016) was employed to derive correlational tractography. The change in MADRS score and the slope were included as explanatory variables. A multiple regression model considered the effects of age and sex on WM.

The connectometry algorithm estimated local connectomes for individual subjects and compiled them into a single local connectome matrix *Y* (Fig. 2E), with each row representing a subject’s local connectome and each column corresponding to common HCP-1065-atlas-derived fiber orientations (Yeh et al., 2016) (Fig. 2D).

Associations between local connectomes and study variables tend to continue a known fiber pathway:

*Y (loacal connectome matrix) = X (matrix with study variables) * B (coefficient matrix)*

To estimate the empirical distribution of B, 5000 bootstrap resampling iterations (default setting) were applied to the row vectors of matrix X. Local connectomes were assessed across varying T-score thresholds (1, 2, 3) and corresponding p-value thresholds adjusted for false discovery rate (FDR) control at 0.05, 0.075, and 0.1 as recommended in the documentation (https://dsi-studio.labsolver.org/doc/gui_cx.html). The most conservative parameter settings were selected for interpretation following visual inspection (Supp. Fig. 1). The deterministic fiber tracking algorithm implemented in DSI Studio (Yeh et al., 2013) was utilized, with a manually delineated cerebellum mask designated as the termination region.

After applying 8000 random permutations to estimate the FDR correction, topology-informed pruning (Yeh, Panesar, et al., 2019) was iterated four times to remove potentially false-positive findings (Yeh, Panesar, et al., 2019). Results were located from the HCP-1065-atlas and interpreted according to the hypotheses.

### Statistical analysis

The remaining statistical analyses were conducted using RStudio (2021.09.0) at significance level of α = 0.05. The change in MADRS score within 6 weeks (delta MADRS score) was calculated as *1 – post-stimulation score / pre-stimulation score*.

Z-standardized values >± 2.5 were used for outlier detection of clinical changes. Median values of RR slopes were chosen for analysis to reduce within-subject variability and were used for connectometry analysis. They were tested for normal distribution using the quantile-quantile plots. Since significant differences between active and sham stimulation have been confirmed, a Pearson correlation between slopes and clinical outcome was performed (Suppl. Fig. 2B).

## 3. Results

A total of 125 patients were initially screened for participation in the study, out of which 92 met the inclusion and exclusion criteria of the trial. Of the 92 included patients, 81 successfully completed the study protocol (11.96% drop-out rate). The reasons for this were varied with medication changes, health problems independent of iTBS and symptom worsening. Of these, one subject only stayed away from the final survey, so the neighboring MADRS value (V5) was imputed here to complete the MRI data.

Two outliers in MADRS with (z =4.02, z = 2.79) were removed, so that a total of 80 subjects were included in the analysis. No serious adverse events were reported during the observation period. The two treatment arms of the cross-over study were comparable (Active-Sham n = 41, Sham-Active n = 39). In the VAS for blinding verification, there was no significant difference between active and sham conditions (t(77) = 1.69, p = 0.09), confirming its effectiveness.

The relative changes in MADRS were almost identical across both arms (M±SD_active-sham_ = 0.25 ± 0.39, M±SD_sham-active_ = 0.34 ± 0.36). There was no significant difference in age (t(78) = 0.72 p = 0.48) and the mean age of the entire sample was 35.89. 33 (41.25 %) of the subjects were females. The concomitant use of medications was assumed to be negligible across crossover groups (*χ*2(1) = 0.15, *p* = 0.70). The severity of symptoms in both groups significantly decreased over the entire observation period (t(78)= 7.03, p < 0.001).

**Table 1:**
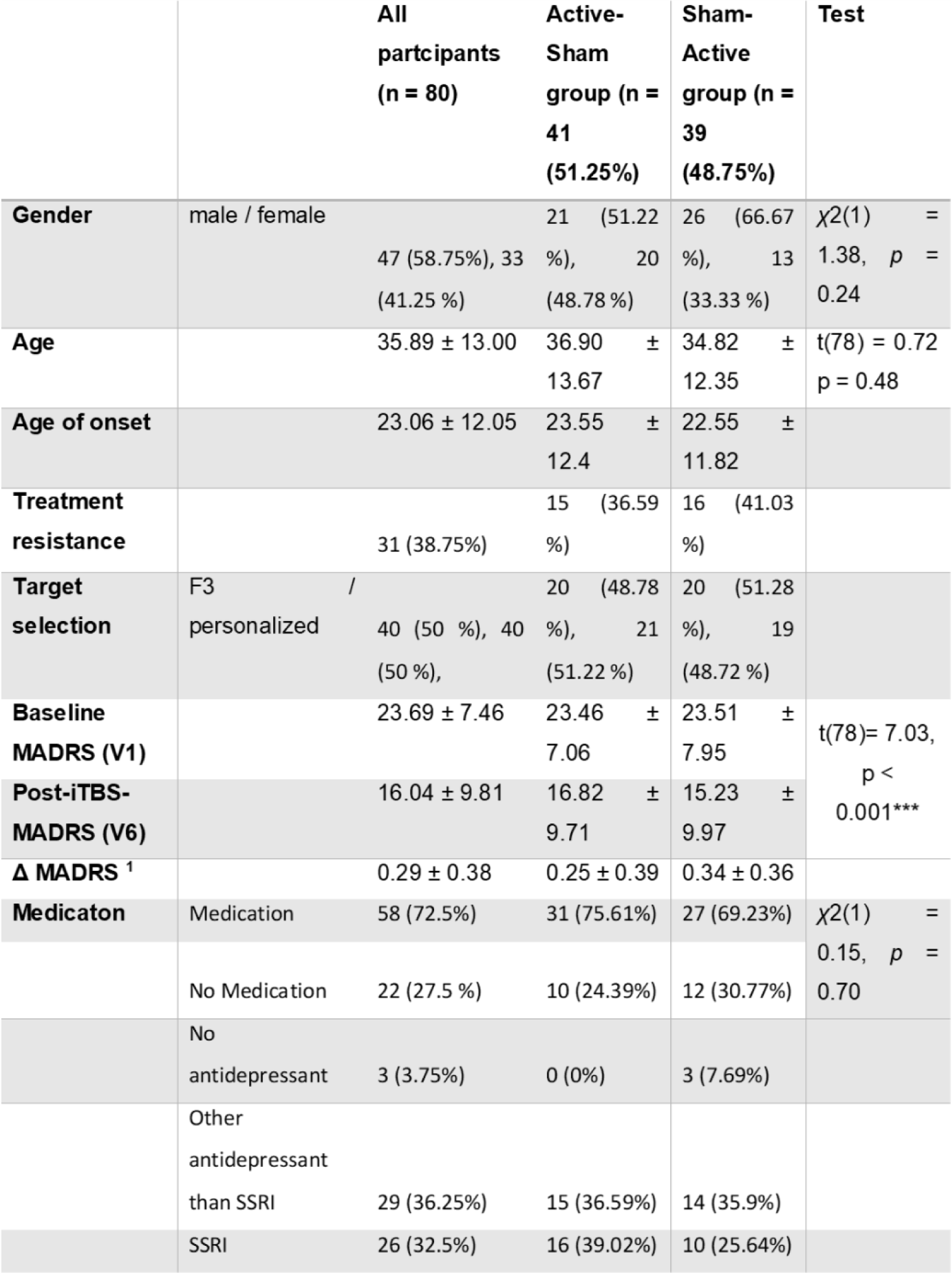
Demographic information and clinically relevant data of the study sample. Delta MADRS^1^ is displayed as relative change (1 – post-stimulation score / pre-stimulation score) and significant test differences are displayed as p < 0.05*, p < 0.01**, p < 0.001***; MADRS: Montgomery-Åsberg depression rating scale.

|  |  | All participants<br>(n = 80) | Active-Sham<br>group (n = 41<br>(51.25%)) | Sham-Active<br>group (n = 39<br>(48.75%)) | Test |
| --- | --- | --- | --- | --- | --- |
| **Gender** | male / female | 47 (58.75%), 33 (41.25 %) | 21 (51.22 %), 20 (48.78 %) | 26 (66.67 %), 13 (33.33 %) | $\chi^2(1) = 1.38, p = 0.24$ |
| **Age** | | 35.89 ± 13.00 | 36.90 ± 13.67 | 34.82 ± 12.35 | $t(78) = 0.72$<br>$p = 0.48$ |
| **Age of onset** |  | 23.06 ± 12.05 | 23.55 ± 12.4 | 22.55 ± 11.82 |  |
| **Treatment resistance** |  | 31 (38.75%) | 15 (36.59 %) | 16 (41.03 %) |  |
| **Target selection** | F3 / personalized | 40 (50 %), 40 (50 %) | 20 (48.78 %), 21 (51.22 %) | 20 (51.28 %), 19 (48.72 %) |  |
| **Baseline MADRS (V1)** | | 23.69 ± 7.46 | 23.46 ± 7.06 | 23.51 ± 7.95 | $t(78) = 7.03,$<br>$p < 0.001^{***}$ |
| **Post-iTBS-MADRS (V6)** |  | 16.04 ± 9.81 | 16.82 ± 9.71 | 15.23 ± 9.97 |  |
| **Δ MADRS <sup>1</sup>** |  | 0.29 ± 0.38 | 0.25 ± 0.39 | 0.34 ± 0.36 |  |
| **Medication** | Medication | 58 (72.5%) | 31 (75.61%) | 27 (69.23%) | $\chi^2(1) = 0.15, p = 0.70$ |
|  | No Medication | 22 (27.5 %) | 10 (24.39%) | 12 (30.77%) |  |
|  | No antidepressant | 3 (3.75%) | 0 (0%) | 3 (7.69%) |  |
|  | Other antidepressant than SSRI | 29 (36.25%) | 15 (36.59%) | 14 (35.9%) |  |
|  | SSRI | 26 (32.5%) | 16 (39.02%) | 10 (25.64%) |  |

The FW-DTI data achieved high data quality (Mean ± SD R^2^_V1_ = 0.65 ± 0.05, Mean R^2^_V4_ = 0.65 ± 0.05). There was extensive overlap in the previously identified relevant tracts in the baseline analysis. Also, significant longitudinal differences emerged in this exploratory analysis. Other tract results apart from the preselected ones are presented in the Supplementary Material.

The primary finding was a positive correlation between changes in FA_T_ within tracts of the right cingulum and ΔMADRS. Significant negative correlations were observed in the bilateral fornices for MD_T_ and RD_T_ measurements. Additionally, the bilateral fornices showed correlations with HR slope, while the right cingulum exhibited a greater number (104) of significant tracts.

In FA_T_, a substantial number of tracts (128) within the right and left cingulum were negatively correlated with HR slope. In contrast, MD_T_ and RD_T_ predominantly showed positive correlations with slope in fibers of the left cingulum (Fig. 3, Tab. 2).

**Figure 3:**
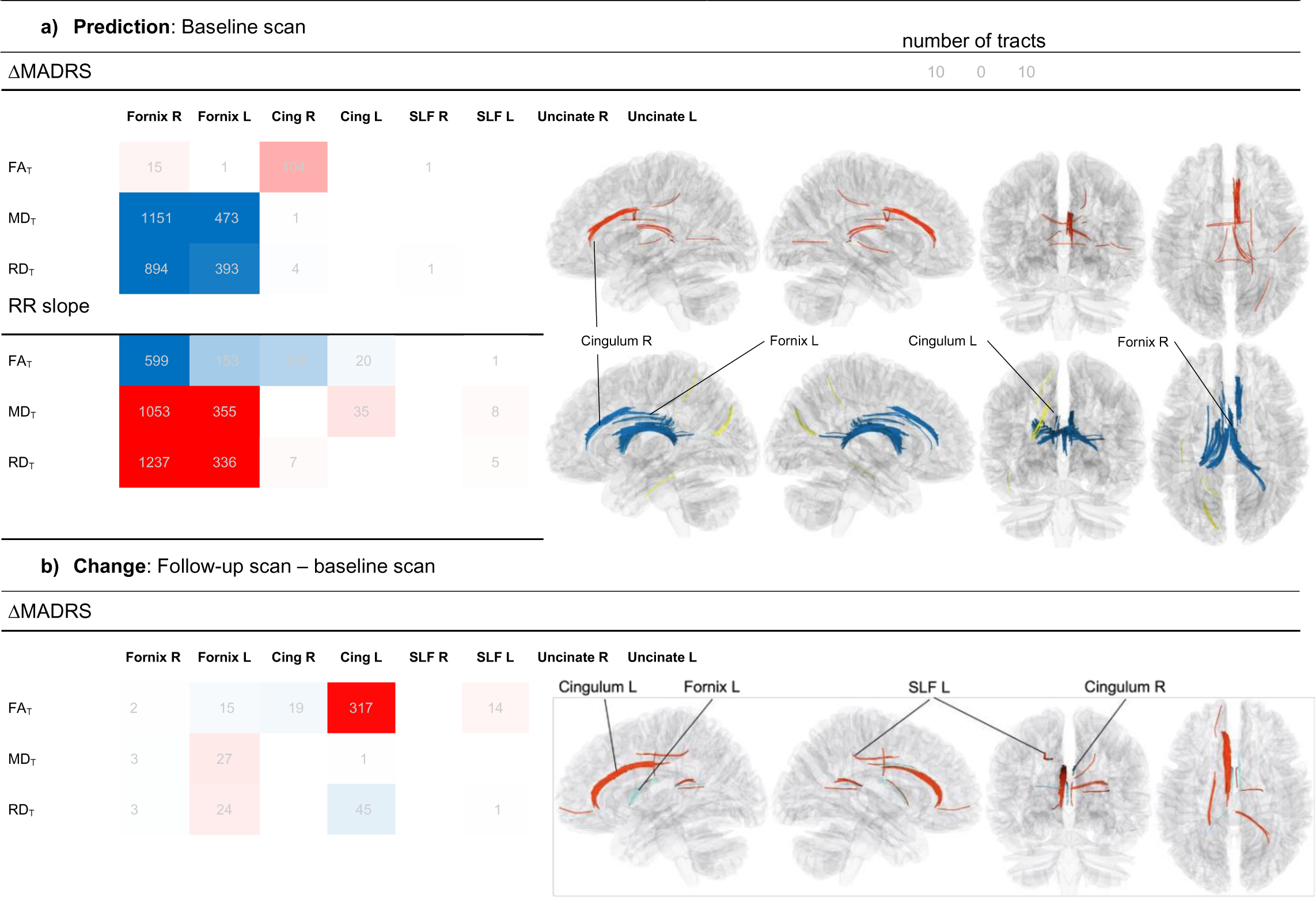
DTI tracts with significant correlation of FW-corrected fractional anisotropy (FA_T_), mean diffusivity (MD T) and radial diffusivity (RD T) with RR slope and delta MADRS. Positive correlations (red) and negative correlations (blue) are shown by number of tracts (grey numbers) in the color scale.

An increase in FA_T_ within the left cingulum significantly correlated with positive clinical outcomes. Similarly, a positive association was identified in a subset of fibers within the SLF. Conversely, a reduction in FA_T_ in a small number of fibers within the right cingulum and left fornix was associated with reductions in MADRS. Additionally, a decrease in RD_T_ in the left cingulum, as well as increases in MD_T_ and RD_T_ in the left fornix, were observed (Fig. 3, Tab. 3).

An overlap was observed in a small number of fibers positively correlated with ΔMADRS and numerous fibers negatively correlated with RR-slope and FA_T_ within the right fornix at baseline. Fibers exhibiting negative correlations of FA_T_ with RR-slope at baseline coincided spatially with regions in the left fornix showing MADRS-associated FA_T_ decreases and MD_T_ increases. In both fornices, these results overlapped with tracts negatively correlated with baseline MADRS and MD_T_.

In the right cingulum, baseline analysis revealed positive associations between FA_T_ and MADRS, as well as tracts with negative correlations between RR-slope and FA_T_, adjacent to regions where FA_T_ showed MADRS-related decreases. Notably, FA_T_-RR-slope tracts in the right cingulum followed a more dorsal trajectory, distinct from the positive FA_T_-MADRS fibers observed in the right fornix. In contrast, findings in the left cingulum showed partial overlap between tracts exhibiting baseline FA_T_-slope correlations and those with FA_T_ increases.

For visualization, only tract numbers >10 are labeled. All displayed tracts have p FDR < 0.05. DTI: Diffusion tensor imaging, FW: Free-water, MADRS: Montgomery-Åsberg depression rating scale, HR: heart rate, RR: Time between two successive ventricular depolarizations.

## 4. Discussion

In this study we investigated therapeutic outcomes in iTBS treatment of MDD, focusing on heart rate (HR) deceleration and white matter (WM) free-water-corrected (_T_) microstructural architecture. iTBS-driven HR deceleration was found to negatively correlate with baseline WM FA_T_ in the right dorsal cingulum and fornix, while symptom alleviation positively correlated with FA_T_ in the right ventral cingulum. Inverted correlations for both symptom alleviation and HR deceleration were found for MD_T_ and RD_T_ predominantly in the fornices (Fig. 3). Longitudinally, FA_T_ increases and RD_T_ decreases in the left cingulum were significantly associated with greater symptom alleviation, evident as early as four weeks.

### Role of the Cingulum and Fornix in Therapeutic Outcomes

The cingulum and fornix emerged as pivotal structures mediating the relationship between HR deceleration and therapeutic outcomes, aligning with their integral roles in the CAN and the pathophysiology of MDD (Liston et al., 2014; Musgrove et al., 2015). The fornix, known for its well-organized white matter tracts, displayed a strong association with positive treatment responses. Specifically, bilateral fornices showed higher fractional anisotropy (FA_T_) alongside lower MD_T_ and RD_T_ values, which correlated with improved MADRS scores. These findings corroborate earlier studies emphasizing the role of intact myelination and white matter integrity in predicting favorable responses to MDD treatments (Fu et al., 2021; He et al., 2021; Ning et al., 2022).

Even so, a more nuanced interpretation is required given the variability in findings. For instance, Korgaonkar et al. (2014) identified lower FA in the fornix as predictive of remission following pharmacotherapy, possibly reflecting distinct mechanisms of neural plasticity across treatment modalities. Nevertheless, the fornix’s involvement in HR modulation is consistent with its integration within the CAN, a network essential for regulating autonomic function and emotional processing (Beissner et al., 2013; Benarroch, 1993; Valenza et al., 2019, 2020).

### Differential Role of the Cingulum in Treatment Response

The cingulum emerged as a critical predictor of therapeutic outcomes, with the right ventral segment showing the strongest association between higher FA_T_ and symptom alleviation. These findings align with prior work identifying elevated FA in the cingulum of treatment responders (Korgaonkar et al., 2014).

Interestingly, within the right cingulum, we observed a decrease in FA_T_ over the observation period in fiber regions that spatially overlapped with tracts where high baseline FA_T_ predicted changes in MADRS. Notably, in the same subregion of the right cingulum, though without spatial overlap, white matter properties demonstrated a negative correlation with heart rate deceleration during iTBS.

This heterogeneity may reflect dynamic neuroplastic processes, including alterations in axonal diameter, shifts in intra- and extra-axonal water content, or regional remodeling driven by iTBS-induced neural activity (Almeida & Lyons, 2017). Such short-term changes in microstructural properties underscore the need for advanced imaging techniques like free-water DTI (FW-DTI) to capture subtle, region-specific alterations that are often missed by conventional DTI metrics (Bergamino et al., 2021; Ning et al., 2022; Pasternak et al., 2015; Tang et al., 2021).

### Mechanistic Insights into Observed White Matter Changes

The observed FA increases in the left cingulum underscore its neuroplastic potential, with changes indicative of myelination and protective remodeling occurring within just four weeks of treatment. These findings align with studies linking FA increases to learning- and stimulation-induced neuroplasticity, where myelination and oligodendrocyte activity were key contributors (Blumenfeld-Katzir et al., 2011; Seewoo et al., 2022; Takeuchi et al., 2010). The capacity of iTBS to promote long-term potentiation (LTP)-like effects further supports this interpretation (Huang et al., 2005).

The FA increases in the left cingulum, coupled with MD_T_ and RD_T_ reductions, point to myeloproliferative processes affecting the axon diameter, consistent with enhanced myelin production and oligodendrocyte differentiation (Almeida & Lyons, 2017; Blumenfeld-Katzir et al., 2011; Sherafat et al., 2012). Conversely, decreases in FA_T_ and increases in MD_T_ and RD_T_ observed in the left fornix and right cingulum may reflect alternative neuroplastic processes, such as axonal branching, neurofilament stain or increased cellular volume, which are often associated with experience-dependent learning (Hofstetter et al., 2013; Taubert et al., 2010).

The rapid microstructural changes induced by iTBS are unlikely to represent uniform myelination alone and may reflect transient shifts in axonal water content or extracellular matrix remodeling. The bi-tensor model used in this study provides a more granular understanding of these dynamic changes. However, multi-shell imaging or model-free reconstruction approaches could further enhance the interpretability of complex structural alterations (Golub et al., 2021; Zhang et al., 2020).

### Limited Predictive Value of the Uncinate Fasciculus and Superior Longitudinal Fasciculus

The uncinate fasciculus and SLF demonstrated limited predictive value for treatment outcomes, consistent with previous findings (Barredo et al., 2019). While a few SLF fibers showed correlations with MADRS reductions or HR deceleration, their overall contribution appeared marginal. This does not suggest irrelevance but rather highlights that their roles in depression and autonomic regulation may involve indirect or compensatory mechanisms that were not detectable in this study.

### Limitations and Future Directions

The study’s reliance on single-shell DTI sequences limits its ability to differentiate between restricted and non-restricted diffusion, reducing the biological specificity of observed changes. Multi-shell imaging or advanced methodologies like generalized q-sampling imaging (GQI) would provide greater insight into underlying neurobiological mechanisms (Yeh et al., 2010). Additionally, the absence of a sham stimulation group and the potential confounding influence of concurrent antidepressant use constrain the causal inferences that can be drawn.

Future research should incorporate longer follow-up periods to capture sustained neuroplastic changes and evaluate the durability of iTBS-induced effects. Combining functional seed-based analyses with connectometry could help elucidate the relationship between structural remodeling and functional connectivity in response to iTBS.

Heart rate deceleration also emerged as a robust marker of treatment efficacy, consistent with its role in autonomic regulation and its anticorrelation with sgACC activity (Dijkstra et al., 2024; Fox et al., 2012; Weigand et al., 2018). The interplay between HR modulation, the frontal-vagal network, and subcortical structures warrants further investigation to refine its utility as a biomarker for personalized iTBS protocols.

### Conclusion

This study highlights the intricate interplay between white matter architecture, heart rate modulation, and clinical outcomes in iTBS treatment for MDD. The cingulum and fornix emerged as critical predictors of therapeutic response, with observed white matter changes reflecting diverse neuroplastic processes, including myelination, axonal remodeling, and dynamic extracellular matrix shifts. While these findings enhance our understanding of iTBS’s structural correlates, further research is needed to refine mechanistic insights and improve the precision of neuroanatomical predictors.

As neuromodulatory treatments continue to evolve, the integration of advanced imaging techniques with autonomic biomarkers holds promise for tailoring interventions to individual neurobiological profiles. Such personalized approaches may improve treatment outcomes for MDD and set a precedent for precision psychiatry in other neuropsychiatric disorders.

## Acknowledgements

We thank the study participants and the research teams of the Laboratory of Systems Neuroscience and Imaging in Psychiatry and the MR-Research in Neurosciences led by PD Dr. Peter Dechent at the University Medical Center Göttingen. We also thank Dr. Fang-Cheng Yeh, MD, PhD at the Department of Neurological Surgery, University of Pittsburgh, for providing continuous technical advice and excellent support.

## Data availability

The data supporting the findings of this study are available on reasonable request from the corresponding author. The data are not publicly available due to privacy or ethical restrictions.

## Supplementary Material

**Supplementary Figure 1:**
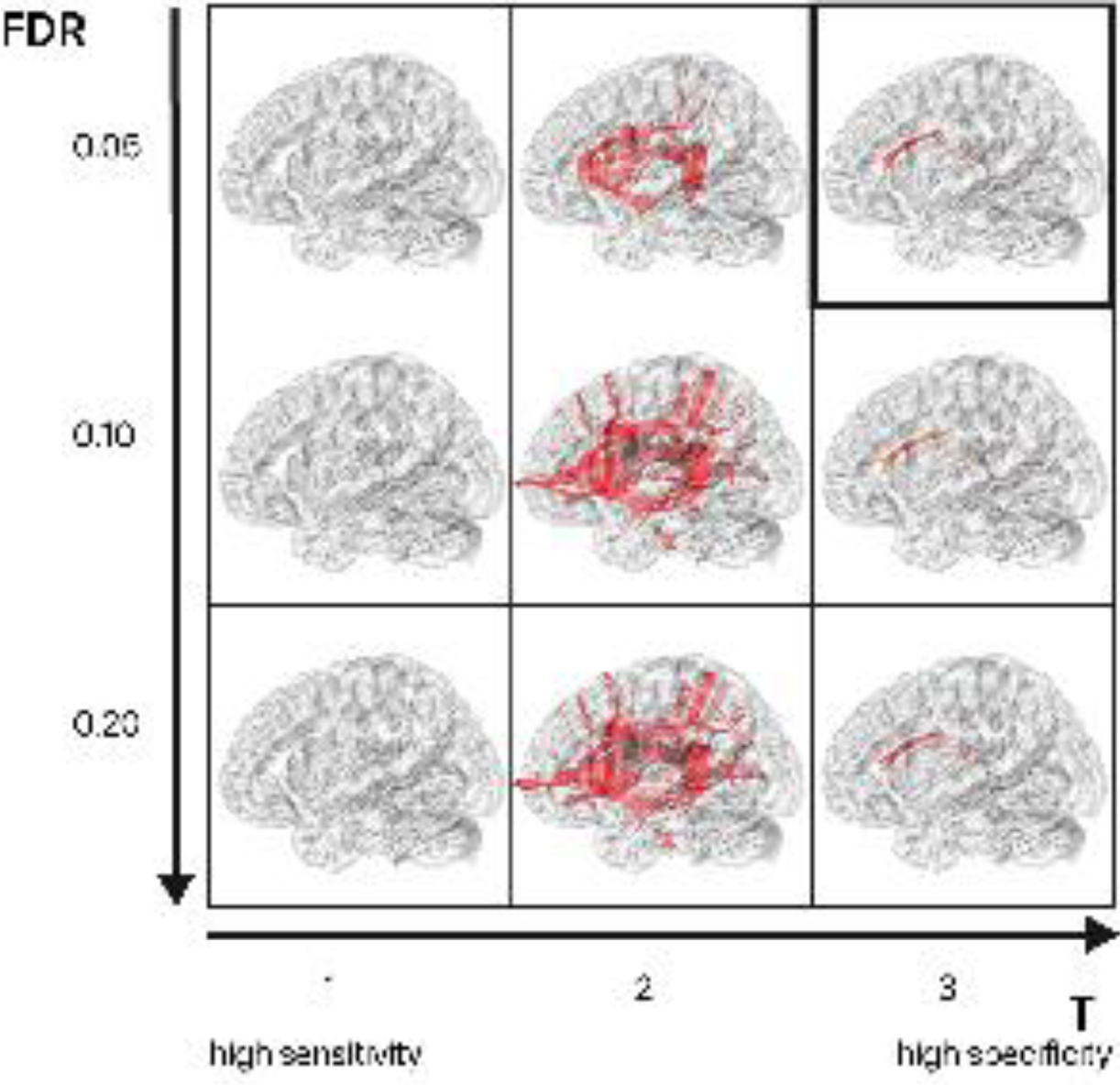
Correlational tractography. Varying t-score thresholds were inspected to select local connectomes (T: 1, 2 and 3) at different significance levels (p FDR < 0.05, < 0.1 and < 0.2) as suggested by the DSI-Studio documentation (https://dsi-studio.labsolver.org/doc/gui_cx.html). The black shape marks the value selection for further analysis in this work

**Supplementary Figure 2:**
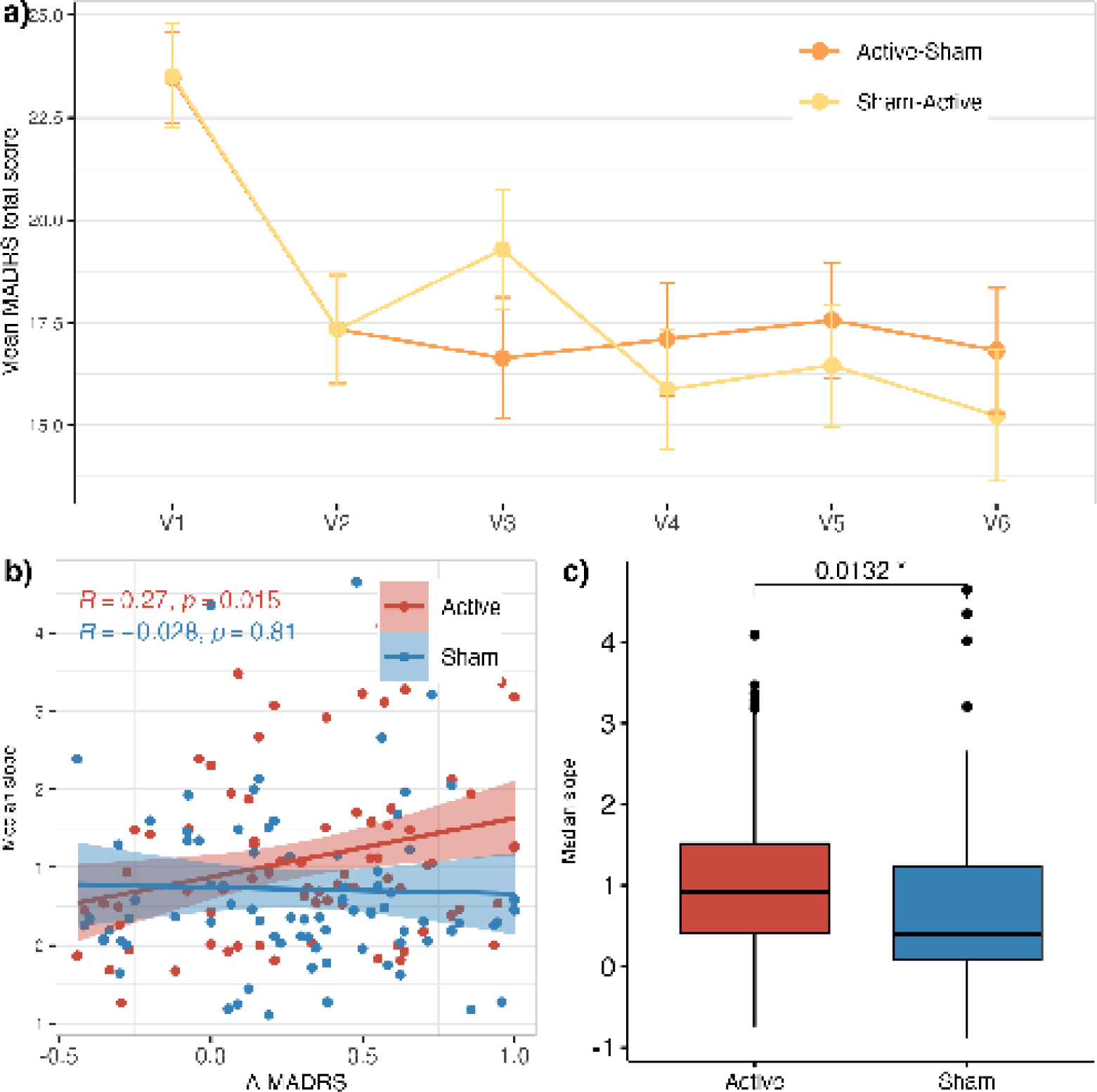
Previously reported clinical findings. a) Montgomery–Åsberg Depression Scale (MADRS) mean total scores in the sham-active and active-sham groups from V1 to V6. b) Pearson correlation between relative delta MADRS and the corresponding slope in RR intervals after 45s of stimulation on the first day of the week. Slope value represents the median of the sessions. c) Comparison of median slope values after 45s of stimulation. Paired t-test t(79) = 2.54, p = 0.0132.

**Supplementary Table 1:** Baseline scans. Recognition of results from correlational tractography based on the HCP1065 tractography atlas.

| V1 deltaMADRS |  |  |
| --- | --- | --- |
| FA<sub>T</sub> deltaMADRS | Positive (132) | 68.1818% Cingulum_Parolfactory_R<br>11.3636% Fornix_R<br>10.6061% Cingulum_Frontal_Parietal_R<br>3.78788% Inferior_Fronto_Occipital_Fasciculus_R<br>1.51515% Corpus_Callosum_Body<br>0.757576% Corpus_Callosum_Forceps_Major |
|  | Negative (0) |  |
| MD<sub>T</sub> deltaMADRS | Positive (0) |  |
|  | Negative (2077) | 55.4165% Fornix_R<br>22.7732% Fornix_L<br>18.1993% Thalamic_Radiation_Superior_R<br>2.02215% Thalamic_Radiation_Posterior_R<br>0.818488% Optic_Radiation_R<br>0.288878% Thalamic_Radiation_Anterior_R<br>0.240732% Corpus_Callosum_Body<br>0.144439% Corpus_Callosum_Forceps_Major<br>0.0481464% Corpus_Callosum_Tapetum |
| RD<sub>T</sub> deltaMADRS | Positive (0) |  |
|  | Negative (1701) | 52.5573% Fornix_R<br>23.1041% Fornix_L<br>20.047% Thalamic_Radiation_Superior_R<br>2.46914% Thalamic_Radiation_Posterior_R<br>0.705467% Optic_Radiation_R<br>0.235156% Thalamic_Radiation_Anterior_R<br>0.176367% Corpus_Callosum_Body<br>0.117578% CNIII_R<br>0.0587889% Dentatorubrothalamic_Tract_R |
| V1 Slope |  |  |
| FA<sub>T</sub> Slope | Positive (9) | 77.7778% Corpus_Callosum_Forceps_Major<br>11.1111% Superior_Longitudinal_Fasciculus1_L |
|  | Negative (991) | 60.444% Fornix_R<br>15.4389% Fornix_L<br>11.2008% Cingulum_Frontal_Parietal_R<br>8.07265% Thalamic_Radiation_Superior_L<br>1.41271% Cingulum_Parolfactory_R<br>1.10999% Cingulum_Frontal_Parietal_L<br>1.00908% Corpus_Callosum_Forceps_Major<br>0.504541% Cingulum_Parolfactory_L<br>0.302725% Cingulum_Frontal_Parahippocampal_R<br>0.100908% Corpus_Callosum_Body |
| MD<sub>T</sub> Slope | Positive (1649) | 63.8569% Fornix_R<br>21.5282% Fornix_L<br>8.91449% Thalamic_Radiation_Superior_L<br>2.00121% Cingulum_Frontal_Parietal_L<br>1.94057% Corpus_Callosum_Forceps_Major<br>0.545785% Thalamic_Radiation_Anterior_L<br>0.303214% Corpus_Callosum_Body<br>0.242571% Superior_Longitudinal_Fasciculus1_L<br>0.121286% Optic_Radiation_R<br>0.0606428% Thalamic_Radiation_Posterior_R |

**Supplementary Table 2:** Delta scans. Recognition of results from correlational tractography based on the HCP1065 tractography atlas.

| V1 V4 deltaMADRS |  |  |
| --- | --- | --- |
| FA deltaMADRS | Positive (342) | 80.9942% Cingulum_Parolfactory_L<br>11.1111% Cingulum_Frontal_Parietal_L<br>4.09357% Superior_Longitudinal_Fasciculus1_L<br>2.04678% Corpus_Callosum_Forceps_Major<br>0.584795% Fornix_R<br>0.292398% Corpus_Callosum_Body |
|  | Negative (35) | 42.8571% Fornix_L<br>40% Cingulum_Parolfactory_R<br>14.2857% Cingulum_Frontal_Parietal_R<br>2.85714% Corpus_Callosum_Tapetum |
| MD deltaMADRS | Positive (105) | 53.3333% Thalamic_Radiation_Superior_R<br>25.7143% Fornix_L<br>16.1905% CNIII_R<br>2.85714% Corpus_Callosum_Forceps_Major<br>0.952381% Middle_Cerebellar_Peduncle |
|  | Negative (78) | 48.7179% Thalamic_Radiation_Anterior_L<br>33.3333% Thalamic_Radiation_Superior_L<br>7.69231% Corpus_Callosum_Forceps_Major<br>3.84615% Fornix_R<br>2.5641% Corpus_Callosum_Tapetum<br>1.28205% Cingulum_Frontal_Parahippocampal_L |
| RD deltaMADRS | Positive (100) | 52% Thalamic_Radiation_Superior_R<br>24% Fornix_L<br>18% CNIII_R<br>2% Reticular_Tract_R<br>1% Middle_Cerebellar_Peduncle |
|  | Negative (144) | 33.3333% Thalamic_Radiation_Superior_L<br>26.3889% Cingulum_Parolfactory_L<br>25% Thalamic_Radiation_Anterior_L<br>3.47222% Corpus_Callosum_Forceps_Major<br>2.08333% Fornix_R<br>1.38889% Reticular_Tract_L<br>0.694444% Superior_Longitudinal_Fasciculus1_L |

